# Dynamic Prediction of SARS-CoV-2 RT-PCR status on Chest Radiographs using Deep Learning Enabled Radiogenomics

**DOI:** 10.1101/2021.01.10.21249370

**Authors:** Wan Hang Keith Chiu, Dmytro Poplavskiy, Sailong Zhang, Philip Leong Ho Yu, Michael D. Kuo

## Abstract

Reverse Transcription-Polymerase Chain Reaction (RT-PCR) is the gold standard for diagnosis of SARS-CoV-2 infection, but requires specialized equipment and reagents and suffers from long turnaround times. While valuable, chest imaging currently only detects COVID-19 pneumonia, but if it can predict actual RT-PCR SARS-CoV-2 status is unknown. Radiogenomics may provide an effective and accurate RT-PCR-based surrogate. We describe a deep learning radiogenomics (DLR) model (RadGen) that predicts a patient's RT-PCR SARS-CoV-2 status solely from their frontal chest radiograph (CXR).

## Brief Introduction

Reverse Transcription-Polymerase Chain Reaction (RT-PCR) is the gold standard for diagnosis of SARS-CoV-2 infection^1^, but requires specialized equipment and reagents and suffers from long turnaround times. While valuable, chest imaging currently only detects COVID-19 pneumonia, but if it can predict actual RT-PCR SARS-CoV-2 status is unknown. Radiogenomics may provide an effective and accurate RT-PCR-based surrogate^2^. We describe a deep learning radiogenomics (DLR) model (RadGen) that predicts a patient’s RT-PCR SARS-CoV-2 status solely from their frontal chest radiograph (CXR).

## Methods

The RadGen architecture, based on *SE-ResNeXt-50-32×4d*, was pretrained on ImageNet and ChestX-ray14 and 28,430 CXR from PadChest, and Kaggle before fine-tuned using CXR from a multinational cohort of RT-PCR tested patients from Hong Kong, GITHUB, SIRM and BIMCV (6,326 images)^3-5^. The model first predicted and selected only frontal CXR images, then predicted a segmentation mask of the cropped lung areas to reduce model fitting to unrelated parts of the image before using the segmented area as input for the RT-PCR SARS-CoV-2 binary classification task. The final prediction score was an ensemble consisting of the average of 4 models.

## Results

RadGen achieved a mean Area Under the ROC curve (AUROC) of 0.959 (95%CI 0.955,0.962), sensitivity of 80.8% (3007/3723) and specificity 95.1% (16206/17033) using a pre-determined 0.4 cutpoint. It distinguishes pre-COVID-19, laboratory confirmed pneumonia from SARS2-CoV-2 cases with a specificity of 89.3% (225/252), and a specificity of 96.4% (106/110) on excluding SARS-CoV-2 infection on patients pre-COVID-19 CXR who later became RT-PCR positive.

The RT-PCR-tested cohort in Hong Kong consisted of 314 positive and 2,471 negative patients from 4 hospitals^3^. The SARS-CoV-2 positive patients had a median of 9 (range 3-24) RT-PCR tests and 3 corresponding (range 1-8) CXR during their hospitalization (range 3-112 days); 90.8% (285/314) had mild/asymptomatic disease. The sensitivity and specificity of RadGen for predicting SARS-CoV-2 infection on initial presenting CXR was 79.5% (225/283) and 85.2% (2105/2471).

RadGen time course analysis by autocorrelation function (ACF) plot, which describes how well RadGen predicts a patient’s RT-PCR SARS-CoV-2 status over the course of their entire SARS-CoV-2 infection period, was performed revealing a peak lag at 2 days for radiogenomic signature manifestation on CXR after initial RT-PCR diagnosis. The per-film false negative rate was 10.0% (26/261) and 9.3% (21/225) within and after 7 days of the first RT-PCR positive test with a false positive rate of 68.1% (32/47) and 11.3% (45/397) within and after one week of achieving RT-PCR confirmed viral clearance (Fig 1).

**Fig. 1.**
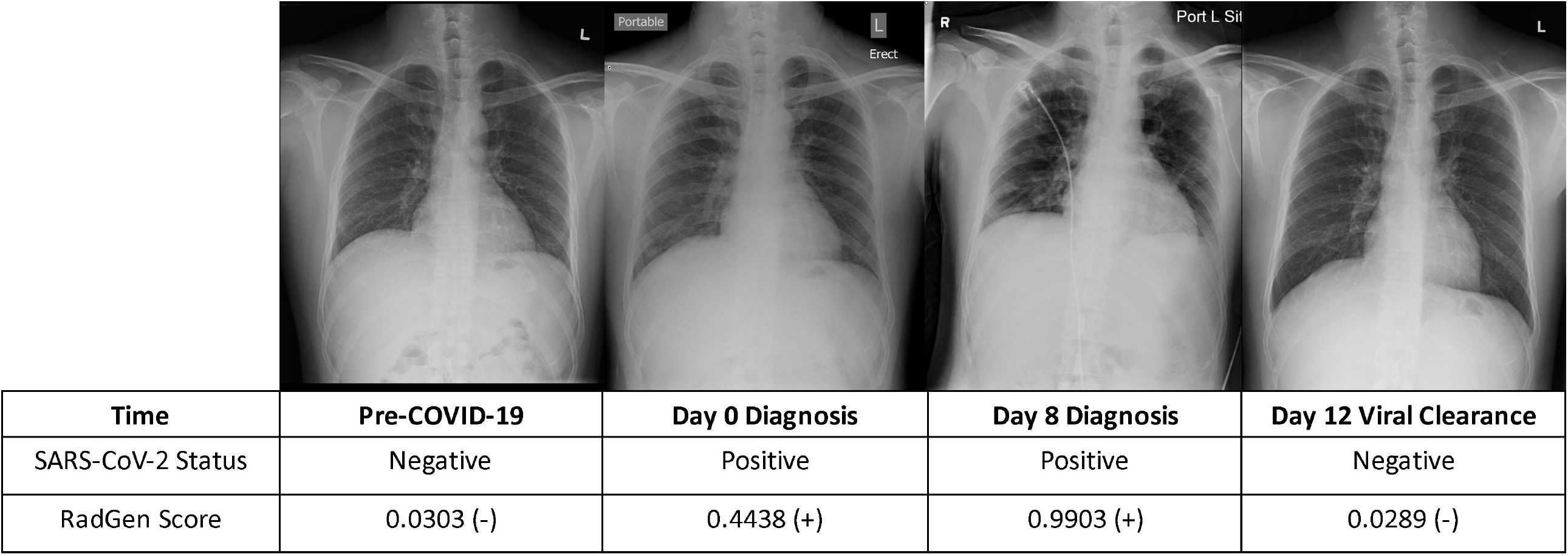
Serial CXR of a SARS-CoV-2 RT-PCR positive male patient in his 40s. RadGen correctly predicted the RT-PCR status of the patient on CXR prior to COVID19 (pre-COVID-19), at initial SARS-CoV-2 RT-PCR confirmed diagnosis (Day 0), during his infection period (Day 8) and upon achieving RT-PCR confirmed viral clearance (Day 12).

## Comment

Leveraging a DLR strategy and a rich body of training datasets including Asian and Western countries (reflecting a diverse set of clinical containment protocols), a wide spectrum of clinical presentations including mild and asymptomatic disease, and a prospectively collected multi-timepoint RT-PCR SARS-CoV-2 positive patient cohort, we generated a DLR model capable of predicting a patient’s RT-PCR status from CXR.

Interestingly, we also show that RadGen can non-invasively ‘track’ RT-PCR SARS-CoV-2 status over the course of their infection, from diagnosis to viral clearance. A time-delayed correlation between RadGen and RT-PCR seen at the time of initial RT-PCR positivity and at the time of achieving RT-PCR viral clearance was observed. This is not unexpected as SARS-CoV-2 genomic dosage changes have been shown to take time to accumulate and be phenotypically reflected on a cellular, organ and systems level. Further, it is known that SARS-CoV-2 RNA can persist long after active infectivity and symptom resolution^6^; thus, that RadGen performs this well, particularly in a mild/asymptomatic cohort, is notable.

In conclusion, the feasibility for DLR models to dynamically track RT-PCR SARS-CoV-2 changes on an individual level significantly expands the scope of radiogenomics.

## Data Availability

Code and models used in this study are available upon reasonable request to the corresponding author and under a collaboration agreement.

## References

1. Kuo MD, Jamshidi N. Behind the Numbers: Decoding Molecular Phenotypes with Radiogenomics—Guiding Principles and Technical Considerations. Radiology. 2014;270(2):320–5.

2. Segal E, Sirlin CB, Ooi C, Adler AS, Gollub J, Chen X, et al. Decoding global gene expression programs in liver cancer by noninvasive imaging. Nature Biotechnology. 2007;25(6):675–80.

3. Chiu WHK, Vardhanabhuti V, Poplavskiy D, Yu PLH, D.R, Yap AYH, et al. Detection of COVID-19 Using Deep Learning Algorithms on Chest Radiographs. Journal of Thoracic Imaging. 2020;Publish Ahead of Print.

4. Iglesia la de Vayá M, Saborit JM, Montell JA, Pertusa A, Bustos A, Cazorla M, et al. BIMCV COVID-19+: a large annotated dataset of RX and CT images from COVID-19 patients2020 June 01, 2020:[arXiv:2006.01174 p.]. Available from: https://ui.adsabs.harvard.edu/abs/2020arXiv200601174I.

5. Bustos A, Pertusa A, Salinas J-M, de la Iglesia-Vayá M. PadChest: A large chest x-ray image dataset with multi-label annotated reports. arXiv e-prints [Internet]. 2019 January 01, 2019:[arXiv:1901.07441 p.]. Available from: https://ui.adsabs.harvard.edu/abs/2019arXiv190107441B.

6. Cevik M, Kuppalli K, Kindrachuk J, Peiris M. Virology, transmission, and pathogenesis of SARS-CoV-2. Bmj. 2020.

