## Supplement for "Dynamic Prediction of SARS-CoV-2 RT-PCR status on Chest Radiographs using Deep Learning Enabled Radiogenomics"

**Supplement Figures**

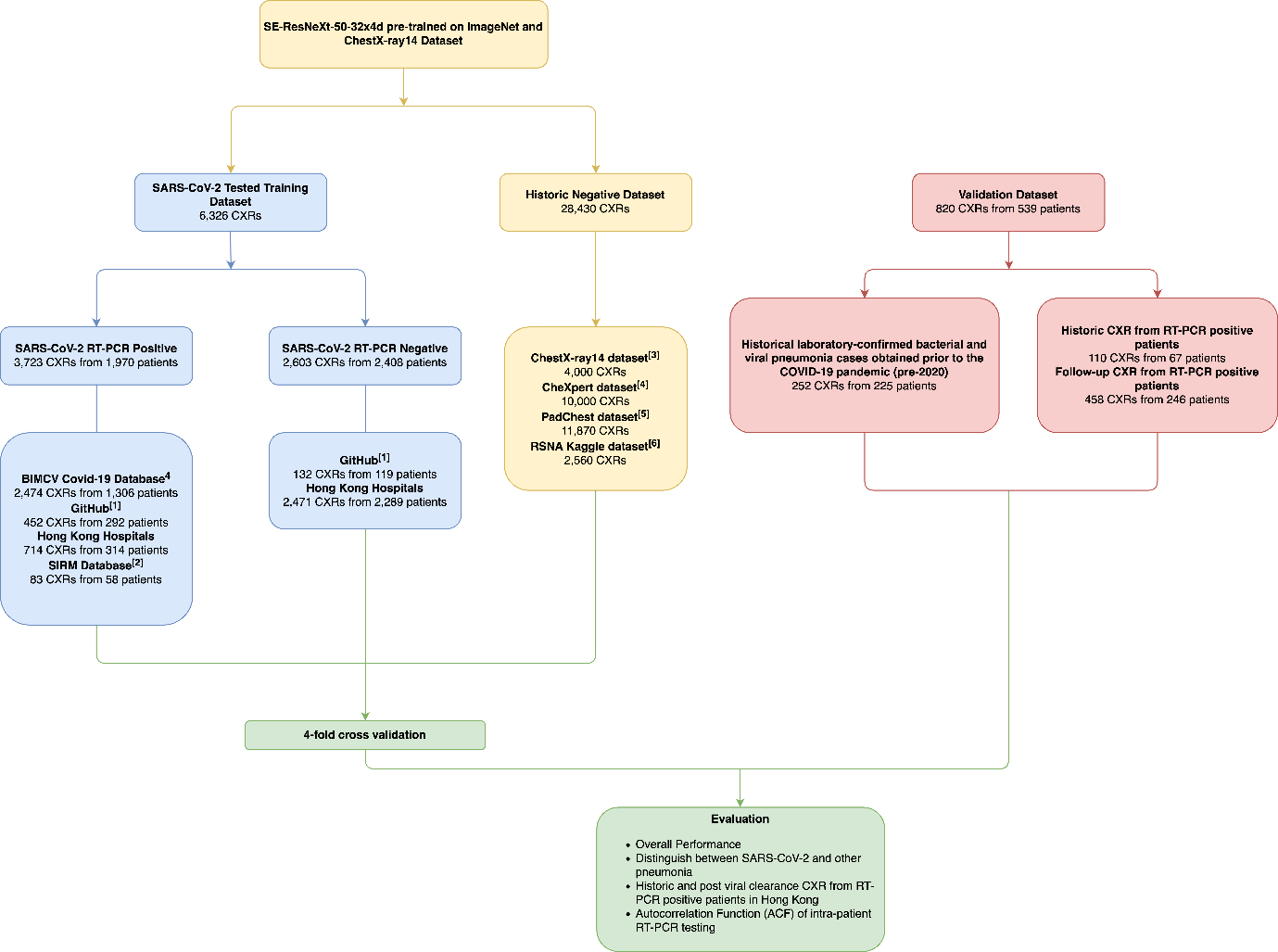

**S1**. Description of datasets and how they were used to construct and validate the RadGen model.

[1] GitHub COVID-19 image data collection (<https://github.com/ieee8023/covid-chestxray-dataset>), Actualmed COVID-19 Chest X-ray Dataset Initiative (<https://github.com/agchung/Actualmed-COVID-chestxray-dataset>), and Figure 1 COVID-19 Chest X-ray Dataset Initiative (<https://github.com/agchung/Figure1-COVID-chestxray-dataset>)

[2] Italian Society of Medical and Interventional Radiology (SIRM) COVID-19 Database (<https://www.sirm.org/en/category/articles/covid-19-database/>)

[3] 4,000 CXR were randomly selected from ChestXray14 dataset of 112,120 CXR for hard negative mining^3^. This dataset was not included for validation.

[4] 10,000 CXR were randomly selected from CheXpert dataset of 224,316 CXR for hard negative mining^3^. This dataset was not included for validation.

[5] 11,870 CXR were randomly selected from the PadChest dataset of over 160,000 CXR and the CXR with the highest false negative prediction scores were used for hard negative mining.

[6] The highest false negative prediction scores of the model from the RSNA pneumonia Detection Challenge dataset on Kaggle was used to perform hard negative mining.

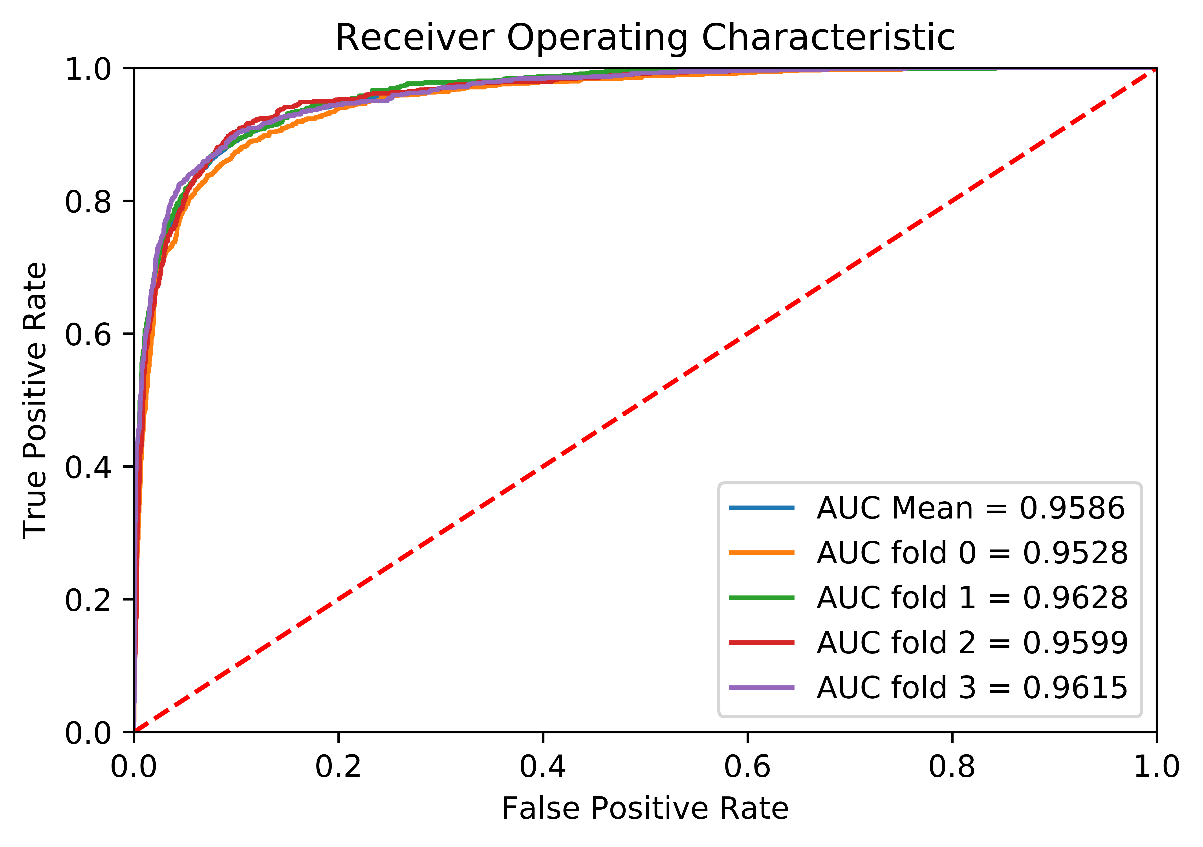

**S2**. Mean and individual Receiver Operating Characteristic (ROC) curves for each validation fold. The model’s operating point of 0.4 was selected based on the maximal Youden index.

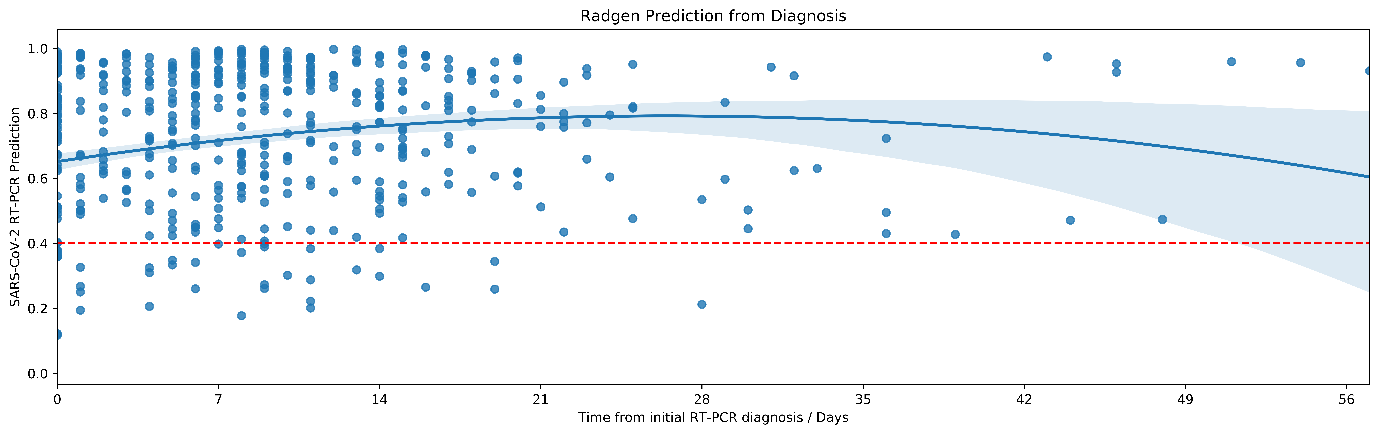

**S3**. Time series profile showing the distribution of prediction scores by RadGen on CXR of all RT-PCR positive patients from the time of their initial diagnosis. Blue area denotes the 95% confidence interval and red dash denotes the RadGen 0.4 cutpoint used to predict RT-PCR binary classification (RT-PCR positive prediction when RadGen score is >0.4).

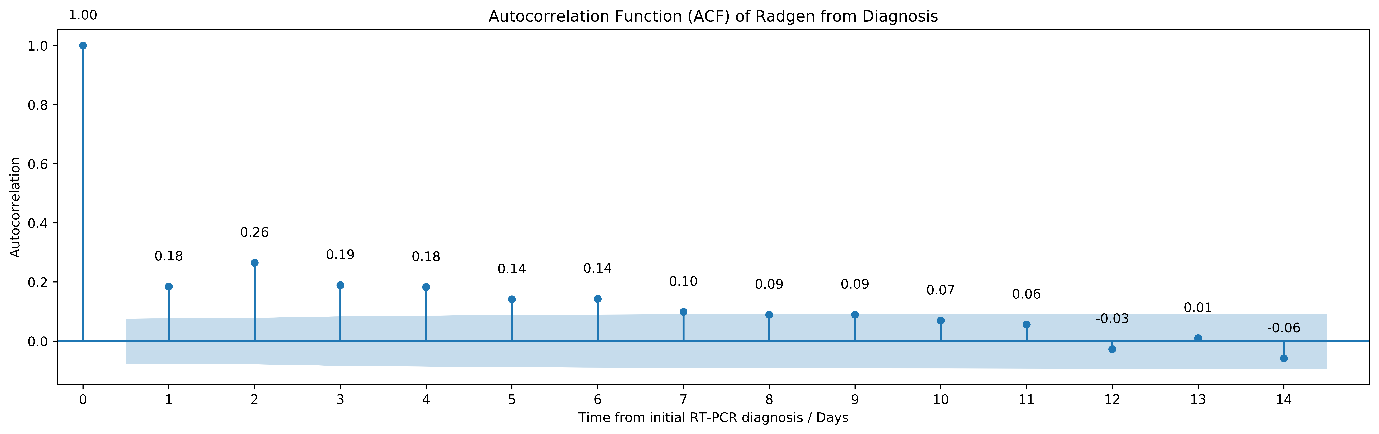

**S4**. Autocorrelation function (ACF) plot of the model demonstrating a time lag over the first 7 days (peak at day 2) between RadGen predicted and corresponding matched RT-PCR status; Blue area denotes 95%CI.
